# Poor evidence for an effect of tecovirimat in shortening recovery time in hospitalized mpox cases from real-world data

**DOI:** 10.1101/2023.02.07.23285617

**Authors:** V Mazzotta, A Cozzi-Lepri, S Lanini, A Mondi, F Carletti, A Tavelli, R Gagliardini, S Vita, C Pinnetti, C Aguglia, P Faccendini, F Colavita, F Faraglia, A Beccacece, J Paulicelli, E Girardi, E Nicastri, F Vaia, F Maggi, A Antinori

## Abstract

**Objectives:** To assess the effectiveness of tecovirimat (TPOXX) for treating mpox in terms of difference in healing time and extent of viral clearance.

**Design:** Emulation of a target trial based on observational data.

**Setting:** Italy

**Participants:** Forty-one men hospitalized for mpox as of September 29^th^, 2022.

**Main outcome measures:** Main outcome was the time to clinical recovery. Secondary outcome was the variation in viral load in the upper respiratory tract (URT) after treatment.

**Results:** The median time from symptoms onset to hospital admission and to initiation of TPOXX was 4 days (IQR 2-6) and 10 days (IQR 8-11), respectively. Fifteen patients completed a course of therapy. No deaths were observed; the overall median healing time was 21 days (IQR 17-26). We found no evidence for a significant improvement in recovery time in treated vs. untreated patients, with an estimated mean of 14.7 days for both groups. A subset of 13 patients had URT samples at T1 (median of 5 days (IQR 3-7) from symptoms onset) and T2 (median 7 days (IQR 7-9) from T1). Overall, mean viral load was 4.65 (0.30) vs. 4.91 (0.35) (log2 scale of cycle threshold) at T1 and T2, respectively. In the unadjusted analysis, variation over T1-T2 was lower in the treated 0.13 log2 (SD=0.53) vs. untreated 0.37 (0.50), although not statistically significant (unpaired t-test p=0.41). After controlling for confounding, there was no evidence for a difference in the potential changes over T1-T2 by treatment arm, and our estimate of the average treatment effect (ATE) was consistent with no difference by treatment group, although with large 95% CI around these estimates.

**Conclusions:** Our analysis seems to exclude a clinically important effect of TPOXX in hospitalized mpox patients when compared to no treatment. These data are one of the valuable currently available sources of evidence to guide treatment decisions in patients hospitalized with TPOXX. Pending more robust data from randomized comparisons, the use of TPOXX should be restricted to the clinical trials setting.

**Trial registration:** “MpoxCohort” observational study protocol: approval number 40z, Register of Non-Covid Trials 2022.

## Introduction

On July 23, 2022, the World Health Organization (WHO) declared mpox to be a Public Health Emergency of International Concern (PHEIC) ^[1]^. Among the 85,159 mpox confirmed cases registered worldwide as of January 29^th^, 2023, only 88 deaths occurred^[2]^, and usually, the disease improved without any antiviral treatment. However, complications leading to hospitalization may occur, and the illness may last for several weeks, during which patients are forced into isolation.

The Centers for Disease Control and Prevention (CDC) suggested to consider mpox treatment in people with severe disease or involvement of anatomic areas, which might result in serious sequelae, or in immunocompromised people or at high risk for severe disease^[3]^. Moreover, the recommended timing for the start of therapy is early after onset.

However, there are currently no proven therapeutics to shorten healing times in mpox. Tecovirimat (TPOXX) was authorized in USA^[4]^ and EU^[5]^ for use against mpox based on promising results from initial studies in animals^[6]^ and evidence of safety in healthy human volunteers^[7]^; TPOXX is also the first choice treatment for mpox suggested by CDC^[3]^. In recent series, oral TPOXX was reported as safe and well tolerated^[8-13]^, no worsening was observed in treated patients, and subjective improvement was reported after a median time of 3 days after treatment. However, the lack of a control group made it difficult to assess treatment effectiveness, and the limited supply did not allow the early use of TPOXX. Furthermore, randomized controlled trials (RTCs) are still ongoing^[14-16]^, and there is an urgent need for robust evidence in order to guide clinical decision-making^[17]^. Nevertheless, the rapidly decreasing number of mpox cases worldwide will probably not allow RCTs to be concluded soon.

Our study aimed to assess the difference in healing time and extent of viral clearance between patients treated and untreated with tecovirimat using observational data to emulate a hypothetical target trial.

## Methods

### Study Population

We included all adult patients with laboratory-confirmed mpox admitted at the Lazzaro Spallanzani National Institute for Infectious Diseases (INMI; Rome, Italy) from May 19^th^ to September 29^th^ and hospitalized for mpox.

Demographic, epidemiological, and clinical characteristics were collected at the time of diagnosis, and diagnostic testing for the mpox virus (MPXV) was performed.

The patients treated with TPOXX received a course of 600 mg twice daily for 14 days: the decision regarding treatments was based on international medical consensus and the availability of drugs.

Participants were followed-up for at least 30 days from symptoms onset until complete clinical recovery or last clinical follow-up.

Data on MPXV viral load in samples collected during the hospitalization from upper respiratory tract (URT), including oropharyngeal swabs and saliva, were recorded. Viral DNA was extracted by the automatic extractor QIAsymphony (Qiagen, Hilden, Germany), and amplified using the real-time PCR method targeting the tumor necrosis factor receptor gene, G2R. MPXV DNA concentration was measured using cycle threshold (Ct) values of the MPXV-specific PCR.

All patients provided written informed consent to participate in the study. The study was approved by the Ethical Committee of the Lazzaro Spallanzani Institute (MpoxCohort protocol: *“Studio di coorte osservazionale monocentrica su soggetti che afferiscono per sospetto clinico o epidemiologico di malattia del vaiolo delle scimmie (Monkeypox)”*; approval number 40z, Register of Non-Covid Trials 2022).

### Statistical analysis

Because the time of symptom onset did not coincide with the date of treatment initiation, to assess the effectiveness of TPOXX, in our target population, we used a counterfactual framework accounting for immortal time bias.

The primary endpoint for the comparison between treated and untreated patients was the time from the date of symptom onset to achieving complete clinical recovery by day 21, defined as the healing of skin and mucosal lesions. Our secondary endpoint was the variation in Ct values in URT from a median of 6 days from the date of symptom onset (T1) to a median of 5 days after T1 (T2).

For the primary endpoint analysis, we aimed to emulate a parallel trial design. The treatment strategies were defined as to start or not to start TPOXX within 10 days from symptoms onset; this interval has been consequently chosen as the grace period in the analysis. The per-protocol effect of TPOXX initiation within 10 days of clinical onset on the primary outcome was quantified by the differences between strategies in (i) not achieving clinical recovery by day 21 (failure) and (ii) restricted mean survival times (survival time difference over a 21-days window) ^[18]^. We assumed that at hospital admission all participants were equally likely to be offered treatment with TPOXX. We created two clones of each participant, with one clone allocated to each strategy, hence doubling the size of our dataset. The study arms in this newly created pseudo-population were, therefore, identical with respect to demographics and clinical characteristics at the time of entering the hospital, thus minimising confounding bias at baseline. In each arm, participants’ follow-up times have been censored when their treatment was no longer compatible with the treatment strategy for the arm (e.g. when there was a deviation from the planned protocol). In our analysis, this occurred for: (i) participants in the no TPOXX arm (control) who received TPOXX within 10 days from symptoms onset and whose follow-up was censored at their time of starting TPOXX and (ii) participants in the treated arm who did not receive TPOXX within 10 days from symptoms onset and whose follow-up was censored at 10 days (including participants who started TPOXX after 10 days). For each participant, clinical recovery by day 21 (if achieved at all) was attributed to the arm in which the participant was still uncensored at the time of the event (i.e., the arm the participant is compliant with). Because there are common causes of the probability of treatment initiation and that of clinical recovery, the artificial censoring introduced by cloning is usually informative. For the comparison of interest, we identified the following potential confounders: age, HIV status, and disease severity, defined as the presence of more than 20 skin lesions and/or involvement of anatomic areas which might result in serious sequelae (e.g., proctitis, pharyngotonsillitis, or ocular involvement). We use inverse probability of censoring weights to control for the cloning-induced informative censoring bias. The 95% Cis were calculated using 100 bootstrap replicates.

In the analysis of the secondary endpoint (Ct variation over time), in order to control for immortal time bias, we used a ‘matching on time’ type of analysis^[19]^. In this approach, participants who initiated TPOXX after hospitalisation were matched to those who did not receive TPOXX and were followed-up for the same amount of time from the date of symptom onset. For example, if a participant started TPOXX 5 days after the date of symptom onset (T1) and had a Ct value measured at T1, the patient was matched to another participant who had not received TPOXX by 5 days who also had a Ct value measured at T1, and both were followed-up from T1 onwards until a second time point (T2) in which a second Ct value was available. We then compared the Ct variation over T1-T2 (in the log2 scale), again by emulating a parallel trial in which TPOXX was the intervention of interest. The average causal effect of TPOXX was estimated using marginal models in which, to control for the effects of age and disease severity, we modelled both the exposure (through inverse probability weighting) and the outcome (via regression) or both (doubly robust). This secondary endpoint analysis was restricted to participants with a Ct value from samples collected from URT available at T1 and T2, and with a T1 value <35. We also conducted a sensitivity analysis controlling for age and HIV status (numerical problems prevented the adjustment for all 3 identified confounders at the same time in this analysis).

## Results

Forty-one hospitalized subjects with mpox were included as of September 29^th^, 2022. Among the 41 patients enrolled, 15 completed a course of TPOXX therapy. In Table 1 main characteristics of patients according to TPOXX exposure were reported. All participants were male, and 95% were self-reported as men who had sex with men. Median age was 35 years (IQR 32-39), and 78% were Caucasian. Only 3 (7.3%) received smallpox vaccine during childhood. Fifteen (36.6%) patients were living with HIV and 17% were HIV negative and received pre-exposure prophylaxis with antiretrovirals; median CD4 cells count in HIV infected was 684 cell/mm^3^ (IQR 471, 884), with no evidence for a difference between TPOXX treated and untreated individuals (Table 1). Overall, 95% of patients had systemic symptoms, and 25 (61%) were classified as having severe disease; 18 (43.9%) had more than 10 mpox cutaneous lesions and main organ disease localizations were proctitis (26.8%), and pharyngotonsillitis (22%). In the original cohort, before cloning took place, the median time from symptoms onset to hospital admission was 4 days (IQR 2-6). The main reasons for hospitalization and treatment were mucosal inflammation and/or superinfection of the lesions and/or management of severe pain due to the lesions. The median time from symptoms onset to initiation of TPOXX was 10 days (IQR 8-11). No deaths were observed, and the overall median time for clinical recovery was 21 days (IQR 17-26).

**Table 1.** Main characteristics of hospitalized patients with monkeypox according to the administration of tecovirimat.

| Characteristics | Treated with Tecovirimat | Untreated | p-value* | Total |
| --- | --- | --- | --- | --- |
|  | N= 19 | N= 22 |  | N= 41 |
| Age, years, *median (IQR)* | 38 (34, 46) | 33 (29, 39) | 0.017 | 35 (31, 39) |
| Smallpox vaccination, *n (%)* | 1 (5.3%) | 2 (9.1%) | 0.643 | 3 (7.3%) |
| Caucasian, *n (%)* | 15 (78.9%) | 17 (77.3%) | 0.898 | 32 (78.0%) |
| MSM<sup>a</sup>, *n (%)* | 18 (94.7%) | 21 (95.5%) | 0.916 | 39 (95.1%) |
| Use of Chemsex, *n (%)* | 0 (0.0%) | 2 (16.7%) | 0.166 | 2 (8.7%) |
| HIV+, *n (%)* | 7 (36.8%) | 8 (36.4%) | 0.975 | 15 (36.6%) |
| CD4 count<sup>b</sup>, *days, median (IQR)* | 714 (471, 1323) | 661 (390, 823) | 0.487 | 684 (471, 884) |
| Use of PrEP<sup>c</sup>, *n (%)* | 3 (15.8%) | 4 (18.2%) | 0.841 | 7 (17.1%) |
| Route of transmission, *n (%)* |  |  | 0.373 |  |
| Recent reported sexual intercourses | 18 (94.7%) | 21 (95.5%) |  | 39 (95.1%) |
| Household | 0 (0.0%) | 1 (4.5%) |  | 1 (2.4%) |
| Systemic symptoms, *n (%)* | 19 (100.0%) | 20 (90.9%) | 0.183 | 39 (95.1%) |
| Number of skin lesions, *n (%)* |  |  | 0.740 |  |
| 0-4 | 6 (31.6%) | 9 (40.9%) |  | 15 (36.6%) |
| 5-10 | 4 (21.1%) | 4 (18.2%) |  | 8 (19.5%) |
| 11-20 | 6 (31.6%) | 4 (18.2%) |  | 10 (24.4%) |
| 21+ | 3 (15.8%) | 5 (22.7%) |  | 8 (19.5%) |
| Pharyngotonsillitis, *n (%)* | 6 (31.6%) | 3 (13.6%) | 0.172 | 9 (22.0%) |
| Proctitis, *n (%)* | 6 (31.6%) | 5 (22.7%) | 0.529 | 11 (26.8%) |
| Ocular Lesions, *n (%)* | 2 (10.5%) | 1 (4.5%) | 0.469 | 3 (7.3%) |
| Severity of disease<sup>d</sup>, *n (%)* | 12 (63.2%) | 13 (59.1%) | 0.793 | 25 (61.0%) |
| Concurrent STIs<sup>e</sup>, *n (%)* | 11 (73.3%) | 9 (69.2%) | 0.814 | 20 (71.4%) |
| Gonorrhoea | 1 (5.3%) | 0 (0.0%) |  | 1 (2.4%) |
| Syphilis | 4 (21.1%) | 4 (18.2%) |  | 8 (19.5%) |
| Other | 6 (31.6%) | 5 (22.7%) |  | 11 (26.8%) |
| Time from onset to hospital admission, *days, median (IQR)* | 5 (2, 6) | 4 (2, 6) | 0.462 | 4 (2, 6) |
| Time from onset to treatment start, *days, median (IQR)* | 10 (8, 11) |  |  | 10 (8, 11) |
| Clinical recovery, *days, median (IQR)* | 23 (18, 29) | 20 (16, 23) | 0.053 | 21 (17, 26) |

<sup>a</sup> Men who have sex with men,
<sup>b</sup> available only in HIV+ participants,
<sup>c</sup> Pre-exposure prophylaxis for HIV
<sup>d</sup> More than 20 skin lesions and/or presence of ocular lesions and/or pharyngotonsillitis and/or proctitis
<sup>e</sup> Sexually transmitted infections;
\* Chi-square or Mann-Whitney U test as appropriate

Our emulation of a parallel trial design analysis showed that, although the risk of 21-day failure was 4.1% lower in participants who were treated with TPOXX vs. those who remained untreated, the 95% CI was large and did not exclude benefit or harm of treatment. Similarly, no significant improvement in recovery time was observed in treated patients, with a mean of 14.7 days estimated for both the treated and untreated groups (Table 2).

**Table 2.** Results of emulation of a parallel trial design with the target population of participants admitted to the hospital for Mpox. It was assumed that treatment initiation was based on the following confounders: age, disease severity, and HIV status. Inverse probability of censoring weights was used to control for the cloning-induced informative censoring bias.

| Original cohort | 21-day failure (%) | 95% CI* |  | Recovery days | 95% CI* |  |
| --- | --- | --- | --- | --- | --- | --- |
| Tecovirimat arm<sup>\$</sup> | 8.3 | 0.0 | 33.3 | 8.4 | 1.9 | 9.2 |
| No Tecovirimat arm | 22.2 | 0.0 | 55.9 | 11.7 | 7.6 | 12.7 |
| *Differences<sup>1</sup>* | -13.9 | -5.2 | +21.0 | -3.3 | -8.8 | +0.4 |
| Emulated cohort | 21 day failure (%) | 95% CI* |  | Recovery days | 95% CI* |  |
| **Kaplan-Meier** |  |  |  |  |  |  |
| Tecovirimat arm | 12.5 | 0.0 | 50.5 | 14.7 | 12.5 | 14.9 |
| No Tecovirimat arm | 15.4 | 0.0 | 31.4 | 14.7 | 12.4 | 14.9 |
| *Differences<sup>2</sup>* | -2.9 | -29.2 | +33.5 | -0.01 | -0.11 | +0.13 |
| **Weighted Kaplan-Meier** |  |  |  |  |  |  |
| Tecovirimat arm | 9.8 | 0.0 | 45.4 | 14.7 | 12.5 | 14.9 |
| No Tecovirimat arm | 13.9 | 0.0 | 29.3 | 14.7 | 12.4 | 14.9 |
| *Differences<sup>3</sup>* | -4.1 | -29.2 | +30.8 | -0.02 | -0.11 | +0.12 |

<sup>\$</sup> Tecovirimat was administered at the dosage of 600 mg twice daily for 14 days
\* The 95% CIs were calculated using 100 bootstrap replicates
<sup>1</sup> These differences are prone to both confounding and immortal-time biases
<sup>2</sup> These differences are prone to informative censoring
<sup>3</sup> These differences account for all types of biases under the assumptions detailed in the methods.

**Table 3.** Potential cycle threshold (Ct) changes (log2 scale) over T1-T2 and Average treatment effect (ATE) from fitting a linear regression model.

| Potential Ct changes (log2 scale) over T1-T2<sup>\$</sup> and ATE<sup>&</sup> from fitting a linear regression model | | | | |
| --- | --- | --- | --- | --- |
|  | Mean<br>in treated with TPOXX<br>(95% CI) | Mean<br>in untreated<br>(95% CI) | ATE<sup>*</sup> (95% CI) | p-value |
| *Treated vs. Untreated* |  |  |  |  |
| IPWs | 0.25 (-0.02, 0.53) | 0.41 (0.08, 0.73) | -0.16 (-0.64, 0.33) | 0.529 |
| Double Robust | 0.38 (0.13, 0.64) | 0.41 (-0.01, 0.84) | -0.03 (-0.58, 0.53) | 0.920 |
| Regression adjustment | 0.38 (0.00, 0.76) | 0.37 (0.01, 0.74) | 0.01 (-0.56, 0.57) | 0.979 |

<sup>\$</sup>T1 was a median of 6 days after symptoms onset; T2 a median of 5 days after T1
<sup>&</sup>Average Treatment Effect
<sup>\*</sup>weighted for age and HIV status

A total of 122 URT samples were collected from 15 treated and 19 untreated patients. Among these, a subset of 13 patients (6 treated and 7 untreated) who had a T1 value <35 and had a second sample at a following time T2, were included in the analysis. Main characteristics of these 13 patients are shown in Supplementary Table 1 and were similar to those of the full cohort. T1 was a median (IQR) of 5 days (3-7) from the date of onset of symptoms, and T2 was 7 days (7-9) from T1. As a consequence of the matching, the timing of T1 and T2 was similar in treated and untreated. Overall, mean Ct values were 25.6 (SD 5.05) and 30.7 (SD 6.6) in the raw scale and 4.65 (0.30) vs. 4.91 (0.35) in the log2 scale at T1 and T2, respectively. The variation over T1-T2 is expressed as the value at T2 minus the value at T1 so positive changes indicate that the participants had a decrease in viral load at T2 (increase in Ct value). In the unadjusted analysis, such increase was lower in the treated 0.13 log2 (0.53) vs. untreated 0.37 (0.50) suggesting poor virological potency of treatment, although not statistically significant (unpaired t-test p=0.41). Results were confirmed by the trial emulation analysis which, after controlling for confounding, showed no evidence for a difference in the potential changes over T1-T2 by treatment arm and our estimate of the average treatment effect (ATE) was consistent with no difference by treatment group, again with large 95% CI around these estimates.

## Discussion

Our trial emulation analysis failed to show a clinical benefit of TPOXX on recovery time or an effect on viral replication in participants who were hospitalized with mpox. The emulation analysis was based on a couple of firm points regarding the choice of the primary outcome, and target population used. At this point in time, given the low case fatality rate of mpox and the required isolation during the presence of active skin lesions, time to recovery could be considered as the primary clinical outcome to be used also in randomized trials^[16]^. our target trial population was restricted to hospitalized patients because the use of TPOXX is currently only suggested for persons with severe mpox^[3]^. In absence of the results from randomized studies our results represent a valuable source of evidence for the effectiveness of TPOXX in this setting.

Similarly to other disease models^[20-22]^, characterizing the early stage of infection as the viral response phase, also for mpox, a timely initiation of TPOXX is believed to be critical for the effectiveness of the treatment. This is because TPOXX is an antiviral and it is reasonable to assume that a prompt reduction in viral replication would lead to quicker clinical recovery.

Our previous, uncontrolled, descriptive data on treated mpox patients showed a progressive decline in MPXV viral load in the course of antiviral treatment^[10]^; nevertheless, in other cohorts^[23-24]^ of untreated patients, viral shedding also occurred mainly during the first two weeks of the disease after which it naturally declined. In our present analysis, there was no evidence for a difference in viral load reduction after an average of 12 days from the date of symptoms onset when comparing treated versus untreated mpox patients after controlling for potential immortal and confounding bias.

Our analysis has a number of limitations. First of all, treatment was not randomly allocated and was initiated a number of days after the date of clinical onset/hospitalization. The delay in treatment initiation was due, in first instance, to the fact that hospitalization typically occurred several days after clinical onset. In addition, once the patient was in the hospital, treatment initiation could have been further delayed by limited drug availability for two main reasons: i) the drug is typically available only for large stocks, and our single-center study has limited sample size; ii) the high cost of the drug. Indeed, in our cohort, the average delay in starting treatment was 10 days after the date of clinical onset. This is consistent with the data of other similar reports showing an average delay of 7-21 days for treatment initiation after hospitalization^[8-10-13]^. This delay in treatment initiation appeared to have led to an artificial beneficial effect of TPOXX in the original cohort analysis, which indeed showed a larger difference between arms for the main outcomes of risk of failure by day 21 and length of clinical recovery. However, immortal time and confounding bias were minimized by our cloning and weighting approach to analysis, which showed a largely attenuated difference by study arm. In addition, by using a fixed grace period of 10 days for treatment initiation, our analysis cannot address the question of whether greater effectiveness of TPOXX might be achieved by, for example, initiating therapy earlier.

Second, our study, especially for the analysis of the virological endpoint, has a very limited sample size and, consequently had low statistical power to detect a difference between arms. Although there seems to be no evidence for a difference by arm from looking at our point estimates, there was a large uncertainty around the estimates, and both benefit and harm could not be excluded with 95% confidence. Third, URT sample collection and storage varied by participants and over time, and therefore the analysis of the virological endpoint could be conducted only on a small subset of the study population. However, the selection appeared to be fairly random, and the characteristics of the included population were similar to those of the whole cohort. Last but not least, as usual when using observational data, we cannot rule out unmeasured confounding bias.

In conclusion, our analysis seems to be able to exclude a clinically important effect of TPOXX in hospitalized mpox patients when compared to no treatment. Despite all the mentioned limitations, our careful analysis of observational data represents one of the valuable current sources of evidence to guide clinical decisions. In light of the fact that most patients seem to recover after a short clinical course of the disease, although a proper cost-effectiveness analysis needs to be conducted, our results, together with the high cost of the drug, appear to suggest a low cost-effectiveness of TPOXX for the treatment of mpox. For these reasons, while awaiting more solid data coming from randomized comparisons, we believe that the use of TPOXX should be restricted to patients enrolled in clinical trials.

## Data Availability

All data produced in the present study are available upon reasonable request to the authors

## Contributors

VM, ACL, and AA contributed to the study concept and design, analysis, and interpretation of data. SL contributed to the critical revision of the manuscript for important intellectual content.

AT, CA, JP, and AB acquired the data. VM and ACL drafted the manuscript. ACL did the statistical analysis. AM, CP, RG, SV, and FF reviewed the manuscript. FC, FCo, and FM were responsible for the virological data. VM and AA are guarantors. All authors reviewed the manuscript.

The corresponding author attests that all listed authors meet authorship criteria and that no others meeting the criteria have been omitted.

## Ethical statement

Data were recorded as part of routine activities and from database of the observational study approved by the Ethical Committee of the Lazzaro Spallanzani Institute (MpoxCohort protocol: *“Studio di coorte osservazionale monocentrica su soggetti che afferiscono per sospetto clinico o epidemiologico di malattia del vaiolo delle scimmie (Monkeypox)”*; approval number 40z, Register of Non-Covid Trials 2022). Data have been analysed anonymously.

## Conflicts of interests

All authors have completed the ICMJE uniform disclosure form at www.icmje.org/disclosure-of-interest/ and declare no conflict of interest for the present study.

The corresponding author (CC) affirms that the manuscript is an honest, accurate, and transparent account of the study being reported, no important aspects of the study have been omitted, and any discrepancies from the study as planned have been explained.

## Acknowledgments

We acknowledge all the participants, nursing staff and the *INMI Mpox Study Group*: Isabella Abbate, Alessandro Agresta, Camilla Aguglia, Alessandra Amendola, Andrea Antinori, Francesco Baldini, Tommaso Ascoli Bartoli, Alessia Beccacece, Rita Bellagamba, Giulia Berno, Aurora Bettini, Nazario Bevilacqua, Licia Bordi, Marta Camici, Priscilla Caputi, Fabrizio Carletti, Rita Casetti, Angela Corpolongo, Stefania Cicalini, Eleonora Cimini, Francesca Colavita, Alessandra D’Abramo, Angela D’Urso, Gabriella De Carli, Patrizia De Marco, Federico De Zottis, Silvia Di Bari, Lavinia Fabeni, Francesca Faraglia, Valeria Ferraioli, Carla Fontana, Federica Forbici, Concetta Maria Fusco, Marisa Fusto, Roberta Gagliardini, Anna Rosa Garbuglia, Saba Gebremeskel Teklé, Maria Letizia Giancola, Giuseppina Giannico, Simona Gili, Emanuela Giombini, Enrico Girardi, Giulia Gramigna, Germana Grassi, Elisabetta Grilli, Susanna Grisetti, Cesare Ernesto Maria Gruber, Eleonora Lalle, Simone Lanini, Daniele Lapa, Gaetano Maffongelli, Fabrizio Maggi, Alessandra Marani, Andrea Mariano, Ilaria Mastrorosa, Giulia Matusali, Silvia Meschi, Valentina Mazzotta, Sabrina Minicucci, Claudia Minosse, Klizia Mizzoni, Martina Moccione, Annalisa Mondi, Vanessa Mondillo, Giorgia Natalini, Nicoletta Orchi, Sandrine Ottou, Jessica Paulicelli, Elisabetta Petrivelli, Maria Maddalena Plazzi, Carmela Pinnetti, Silvia Pittalis, Gianluca Prota, Vincenzo Puro, Silvia Rosati, Alberto Rossi, Gabriella Rozera, Marika Rubino, Martina Rueca, Laura Scorzolini, Eliana Specchiarello, Maria Virginia Tomassi, Massimo Tempestilli, Francesco Vaia, Francesco Vairo, Beatrice Valli, Alessandra Vergori, Serena Vita.

