## Supplemental table 1 for "Poor evidence for an effect of tecovirimat in shortening recovery time in hospitalized mpox cases from real-world data"

|  | **Treatment with TPOXX** | | | |
| --- | --- | --- | --- | --- |
| **Characteristics** | **Treated** | **Untreated** | **p-value** | **Total** |
|  | N= 6 | N= 7 |  | N= 13 |
| ***Age, years*** |  |  | 0.474 |  |
| Median (IQR) | 38 (35, 45) | 39 (23, 42) |  | 38 (33, 42) |
| ***Smallpox vaccination, n(%)*** | 0 (0.0%) | 1 (14.3%) | 0.355 | 1 (7.7%) |
| ***Caucasian, n(%)*** | 5 (83.3%) | 5 (71.4%) | 0.626 | 10 (76.9%) |
| ***MSM, n(%)*** | 6 (100.0%) | 7 (100.0%) |  | 13 (100.0%) |
| ***Use of Chemsex, n(%)*** | 0 (0.0%) | 2 (40.0%) | 0.237 | 2 (25.0%) |
| ***HIV+, n(%)*** | 3 (50.0%) | 3 (42.9%) | 0.805 | 6 (46.2%) |
| ***Use of PrEP, n(%)*** | 2 (33.3%) | 2 (28.6%) | 0.859 | 4 (30.8%) |
| ***Number of lesions*** |  |  | 0.288 |  |
| 0-4 | 3 (50.0%) | 1 (14.3%) |  | 4 (30.8%) |
| 5-10 | 1 (16.7%) | 0 (0.0%) |  | 1 (7.7%) |
| 11-20 | 1 (16.7%) | 2 (28.6%) |  | 3 (23.1%) |
| 21+ | 1 (16.7%) | 4 (57.1%) |  | 5 (38.5%) |
| ***Pharyngotonsillitis, n(%)*** | 2 (33.3%) | 1 (14.3%) | 0.435 | 3 (23.1%) |
| ***Proctitis, n(%)*** | 2 (33.3%) | 1 (14.3%) | 0.435 | 3 (23.1%) |
| ***Ocular Lesions, n(%)*** | 0 (0.0%) | 0 (0.0%) |  | 0 (0.0%) |
| ***Systemic symptoms, n(%)*** | 6 (100.0%) | 5 (71.4%) | 0.171 | 11 (84.6%) |
| ***Concurrent STI, n(%)*** | 5 (100.0%) | 1 (33.3%) | 0.049 | 6 (75.0%) |
| Gonorrhea | 1 (16.7%) | 0 (0.0%) |  | 1 (7.7%) |
| Syphilis | 1 (16.7%) | 1 (14.3%) |  | 2 (15.4%) |
| HPV | 0 (0.0%) | 0 (0.0%) |  | 0 (0.0%) |
| Other | 3 (50.0%) | 0 (0.0%) |  | 3 (23.1%) |
| ***Time from onset to hospital admission, days*** |  |  | 0.942 |  |
| Median (IQR) | 6 (2, 6) | 5 (4, 6) |  | 5 (4, 6) |
| ***Clinical recovery, days*** |  |  | 0.010 |  |
| Median (IQR) | 30 (25, 32) | 17 (16, 21) |  | 21 (17, 29) |
| ***CD4 count^$^, days*** |  |  | 0.275 |  |
| Median (IQR) | 828 (714, 1323) | 637 (232, 884) |  | 771 (637, 884) |
| ***Route of transmission*** |  |  |  |  |
| Recent sexual intercourse | 6 (100.0%) | 7 (100.0%) |  | 13 (100.0%) |
| Household | 0 (0.0%) | 0 (0.0%) |  | 0 (0.0%) |
| ***Time from onset to treatment start, days*** |  |  |  |  |
| Median (IQR) | 11 (9, 11) |  |  | 11 (9, 11) |
| ***Severity of disease^£^, n(%)*** | 4 (66.7%) | 5 (71.4%) | 0.859 | 9 (69.2%) |
| ^*^Chi-square or Mann Whitney U test as appropriate | | | | |
| ^$^available only in HIV+ participants | | | | |
| ^£^More than 20 lesions, Ocular lesions, Pharyngotonsillitis or Proctiti. | | | | |
